# Integrated Clinical Decision Support for Empiric Antibiotic Selection in Sepsis: A Protocol For A Cluster Randomized Cross-Over Trial (IDEAS-CRXO)

**DOI:** 10.64898/2026.02.04.26343404

**Authors:** Derek R. MacFadden, Marion Elligsen, Christopher Graham, Akhil Garg, Caroline Nott, Kit Chan, Rosemary Zvonar, Linda Chou, Kathryn N. Suh, Monica Taljaard, Cory E. Goldstein, Nick Daneman

## Abstract

**Introduction:** As antibiotic resistant organisms and infections continue to proliferate globally, it becomes increasingly difficult to select empiric antibiotic therapy, particularly in patients who stand to benefit from early adequate treatment. The inappropriate treatment of suspected infection, including sepsis, can be both too narrow and too broad. There is a need to optimize and tailor selection of antibiotic therapy to each patient, such that those at risk for infections due to antibiotic resistant organisms receive broader therapy, and those that are not at risk receive narrower antibiotic therapy. By helping clinicians select the right antibiotic for each individual patient we can potentially reduce unnecessarily broad antibiotic prescribing while preserving (and potentially improving) adequacy of treatment. Individualized clinical prediction models and decision support interventions are promising approaches that can meet these needs by better defining patient risk for antibiotic resistant or susceptible infections, but rigorous prospective evaluations are needed to confirm their utility.

**Methods:** We propose a two-period two-sequence non-blinded cross-sectional cluster randomized cross-over trial of an individualized antibiotic prescribing decision support intervention, administered by antibiotic stewardship pharmacists, for providers treating hospitalized patients with suspected infection including sepsis. The trial is taking place at 3 hospitals in Ontario, Canada, with clusters defined as medical, surgical, and critical care services. Eligible patients are adults with suspected infection characterized by initiation of a broad-spectrum antibiotic and recent collection of blood cultures. This decision support intervention will be based on a combination of proven decision heuristics (for Gram-positive organisms), modelled predicted susceptibilities (for Gram-negative organisms), and well-defined syndromes, that are individualized to the patient. Our primary outcome will be whether or not patients are de-escalated from their initial empiric regimen at 48 hours. The target sample size is 18 clusters with anticipated data from 1,440 patients, designed to achieve 80% power to detect an absolute increase in de-escalation at 48 hours of *7.*5%. Secondary outcomes include adequacy of therapy, process measures, and safety.

**Analysis:** The primary and secondary outcomes will be analyzed at the patient-level using generalized linear mixed effects regression with fixed effects for treatment and period to account for the cross-over design, as well as fixed effects for the stratification factors and *a priori* patient risk factors to improve power and efficiency. Cluster and cluster-period random effects will be included to account for within-period and between-period intracluster correlation and the Kenward-Roger correction will be applied to minimize small sample bias. The primary effect estimate will be presented as a risk ratio with 95% confidence interval.

**Ethics and Dissemination:** This trial has research ethics board approval at all participating sites and adheres to the Ottawa Statement ethics guidelines. The study is registered with clinicaltrials.gov, and the results will be published open access in a peer-reviewed journal.

**Registration:** Clinicaltrials.gov, NCT06103500.

**STRENGTHS AND LIMITATIONS:**

- IDEAS-CRXO is a cluster randomized cross-over trial of a pharmacist administered decision support intervention for antibiotic prescribing in patients with suspected infection.
- The intervention is based upon a generalizable algorithm incorporating Gram-positive heuristics, Gram-negative modelled susceptibility, and local well-defined syndrome treatment guidelines.
- The primary outcome is a relevant measure for antibiotic stewardship - antibiotic de-escalation from the initial empiric index regimen at 48 hours from receipt of antibiotics. Secondary outcomes are focused on other clinical, process, and safety measures.
- This rigorous cluster randomized cross-over trial will be conducted with 18 clusters (across three institutions) which will achieve 80% power in a two-period two-sequence design to detect a clinically important absolute difference of 7.5% in the primary outcome at a 5% two-sided significance level.
- This study could be impacted by treatment contamination, however the cluster design will limit this potential source of bias.

## INTRODUCTION

### The Threat of Antimicrobial Resistance

In the last 10 years, antibiotic resistance and antibiotic use in humans have emerged as major public health issues globally. These issues have been the subject of high-level meetings at the United Nations, resulting in action plans being developed by many middle and high-income countries and national and international bodies including the Public Health Agency of Canada and the World Health Organization ^1–3^. It is estimated that by 2050 global mortality due to antimicrobial resistance may outpace deaths due to cancer, and that the global economy will be significantly impacted^2^. A recent landmark study indicates that over 1 million attributable deaths are occurring annually ^4^, and there is an urgent need to improve antibiotic prescribing while not compromising patient outcomes. Suspected infection, including sepsis, is one of the major indications for antibiotic prescribing in hospitals. Individualized prescribing may reconcile the need to optimize prescribing to improve patient outcomes and the need to minimize antibiotic pressure driving antimicrobial resistance.

### Improving Antibiotic Prescribing Can Help Reduce Antibiotic Resistance and Potentially Improve Empiric Antibiotic Coverage

Antibiotic prescribing is felt to be a major driver of antibiotic resistance, both in hospitals and outpatient settings^1,2,4^. Antibiotic stewardship principles aim to support prescribers in choosing the ‘right’ antibiotic at the ‘right’ time, though they are most commonly focused on reducing inappropriate prescribing^1,5,6^. The ideal decision-making approach is to differentiate the subset of patients that require broad-spectrum antibiotics from those that do not^6–8^. This allows for focused prescribing and for a potential reduction in the spectrum of antibiotic use, while maintaining/improving adequacy of treatment. Using model and heuristic-based algorithms for antibiotic decision support has the potential to help providers better determine a patient’s risk for antibiotic resistance and prescribe accordingly^5,9^.

### Existing Evidence for Clinical Decision Support Tools in Infection

Increasingly, computerized clinical decision support tools are recognized for their ability to improve patient outcomes. A recent meta-analysis indicated that these systems can drive positive changes in processes of care^10^. With the growing use of electronic medical records^11,12^, including adoption of versatile interface standards^13^, computerized decision support tools are increasingly becoming implementable with ease and their use is becoming part of standard practice for various syndromes^12^. For the treatment of infectious diseases with antibiotics, a recent systematic review showed a number of interventions that have been studied from simple guideline feedback to complex probabilistic frameworks ^14^. Some of these have explored decision support for inpatients with suspected infection though they have not considered individual risk factors for resistance^15^. A small number of randomized trials of antibiotic decision support have been performed and have focused on helping to guide admission and discharge disposition^16^, the need for early treatment^17^, and risk of decompensation^18^. However, none of them described a clinical decision support tool that aided providers in the selection of antimicrobial therapy in suspected infection including sepsis. Recently the INSPIRE cluster randomized trial evaluated embedded prompts during order entry to inform prescribers of potential antibiotic resistant organism risk, and this helped reduce broad-spectrum antibiotic use^19^. While there is an overall paucity of experimental data, quasi-experimental studies do exist on the use of clinical decision support in sepsis. We previously conducted two single-centre non-randomized studies (IDEAS) showing how patients’ previous culture results can be used to provide guidance regarding the potential individual risk of infections from antibiotic resistant organisms^5,20^. These studies demonstrated that a pharmacist-led audit and feedback intervention improved (prior culture) concordant antibiotic therapy and reduced unnecessary vancomycin use in patients with suspected infection, and de-escalated empiric antibiotic therapy while maintaining or shortening time to adequate therapy in patients with Gram-negative bacteremia^5,20^.

### The Need for a Randomized Trial

There is clearly a need to improve antibiotic prescribing in sepsis, and individualized clinical decision support tools represent an innovative and potentially effective solution. Preliminary evidence from quasi-experimental studies, as well as a general absence of randomized studies on the subject, demonstrate the need for a randomized controlled trial to evaluate a clinical decision support intervention for the treatment of sepsis.

### Purpose of the Trial

The aim of the trial is to determine whether an early stewardship led clinical decision support intervention can improve antibiotic de-escalation in patients with sepsis while maintaining or improving adequacy of antibiotic coverage.

### Study Hypotheses

We can use an innovative antibiotic prescribing decision support intervention for providers empirically treating suspected infection, including sepsis, to drive de-escalation of antibiotic therapy, while maintaining adequacy of therapy.

### Objectives

Our objectives are to determine whether a clinical decision support intervention in suspected infection, including sepsis, will:

1. Improve early antibiotic de-escalation.
2. Improve time to adequate antibiotic therapy.
3. Improve other clinical outcomes including antibiotic spectrum at completion of therapy, hospital mortality, hospital length of stay, extent of antibiotic de-escalation, *Clostridioides difficile* infection (CDI) at 90 days from index, requirement for dialysis during admission, and total days of antibiotic therapy. We also seek to:
4. Describe safety measures associated with a clinical decision support intervention, including escalation of antibiotic therapy, and a composite endpoint of mortality or ICU admission at day 7 from receipt of antibiotic therapy.

## METHODS AND ANALYSES

### Trial Design

This is a two-period two-sequence cross-sectional cluster randomized cross-over trial of a clinical decision support intervention versus usual care for patients with suspected infection, including sepsis, with the intent of supporting early de-escalation of empiric antibiotic therapy. Cluster randomization will: (1) help prevent contamination within services; (2) facilitate delivery of the intervention (which is provider focused) in an efficient/effective manner; and (3) will accommodate the presence of trainees which are directly involved in prescribing and associated (by definition) with staff physicians/services. A cross-over design was chosen to improve power and efficiency and to promote a balance in patient case mix across different treatment conditions. Clusters are defined as medical, surgical, and critical care services within participating hospitals. All 18 participating clusters (6 at each of 3 institutions) will be randomized to one of two sequences. To control for period effects, institutions will be started either simultaneously or in short sequence, where possible. There will be one 3-month intervention and one 3-month control period in every sequence for a total study duration of 6-months (Table 1). At each institution, there will be two critical care clusters, two medical clusters, and two surgical clusters. Separate but related services will be contained within a specific cluster. The services comprising these clusters will be grouped to achieve balancing of expected eligible patient sizes as well as selected at the discretion of the site investigator. Some specialty services will be intentionally excluded, e.g. obstetrical/gynecology given potential for high frequency of excluded patients. See Table 1 below for the planned study randomization and cross-over design.

### Study Population

The trial will include 18 service groups (clusters) at three participating large tertiary care hospitals in Ontario, Canada. From these services, spanning medical to surgical to critical care, prescribing advice will be provided to physicians treating adult inpatients with suspected infection, including sepsis.

### Eligibility Criteria

Eligible patients for prescribing advice must meet the following inclusion/exclusion criteria.

Inclusion:

1. Admitted
2. Age >18 years of age
3. Newly started (within 24 hours of assessment for eligibility) on at least one of the following index antibiotic(s):

I. Vancomycin (by intravenous route)
II. Linezolid
III. Daptomycin
IV. Clindamycin
V. Cefazolin
VI. Cloxacillin
VII. Ceftriaxone
VIII. Ceftazidime
IX. Piperacillin-Tazobactam
X. Meropenem (or Imipenem or Ertapenem)
XI. Ciprofloxacin
4. Blood cultures ordered (within 12 hours before or after initiation of index antibiotics).

Exclusion:

1. Pregnancy/breastfeeding
2. Documented end-of-life (palliative) care and will not be receiving ongoing antibiotic treatment.
3. Previously enrolled in the trial.
4. Positive clinical culture results (those with speciation) for the index infection or blood cultures with Gram-positives identified within 72 hours prior to assessment. (Other cultures that are positive with a Gram-stain result but not speciation will not be an exclusion criterion).
5. Positive explanatory molecular test (e.g. Legionella urinary antigen test, SARS-CoV-2 testing) within 72 hours prior to assessment.
6. Receipt of antimicrobials (not chronic suppression or prophylaxis) in the prior 24-72 hours (except if started in the outpatient setting or emergency department prior to admission in the 24-72 hours).
7. Index prescription is a continuation of an antibiotic given for suppressive chronic therapy or long-standing treatment of an established infection.
8. Index antibiotics are peri-operative only or ordered for <24 hours.
9. Cystic fibrosis.
10. Known to be enrolled in another trial that dictates antimicrobial selection.
11. Not eligible for any of the intervention algorithms (See supplemental figures).

The start date and time of antibiotics will define the index event. This will be considered the administered date and time or best measure according to study site. Initial antibiotics given in the emergency department prior to admission/index event will not be part of the 24-72 hour exclusion window given variation in capture of this prescribing across hospitals.

A patient will be considered included in the study when they have been screened and reviewed by the study pharmacists (via electronic medical records) and determined to be eligible according to the noted eligibility criteria. If there is a change in intervention eligibility after this determination, the patient will still be included for outcomes measurement but no recommendation will be provided (the provider gives usual care). This will ensure comparability between control and intervention groups that might otherwise be compromised by processes that differ between control and intervention groups (e.g. communicating with the treating team and being informed that antibiotic treatment was stopped). (See eligibility steps in Supplemental Materials).

Prior antibiotics with minimal systemic absorption or relevance to the intervention drugs will be excluded from consideration of ‘prior antibiotic exclusion’ (e.g. rifaximin, metronidazole, oral vancomycin). Eligible antibiotics for inclusion do not include those administered via the intraperitoneal route.

### The Intervention

The planned intervention consists of a pharmacist-facilitated clinical decision support intervention, where pharmacists make recommendations on empiric antibiotic selection to hospital providers based upon a previously developed ‘Early-IDEAS’ algorithm (Supplemental Figures 1-3)^9^. For patients who are being treated empirically with broad spectrum antibiotics for suspected sepsis within an intervention period, a stewardship pharmacist will review their prior microbiology and epidemiologic risk factors for antibiotic resistance and will recommend either changing antimicrobial therapy (to a narrower, or sometimes broader, antibiotic regimen), or not changing therapy based on the applicable Early-IDEAS algorithm. As with the Early-IDEAS algorithm, some well-defined syndromes (e.g., community-acquired pneumonia, meningitis, cellulitis, post-neurosurgical infection, chronic obstructive pulmonary disease exacerbation, head and neck infection, endocarditis, sinusitis, endocarditis), will follow institutional guidelines. This approach supports the prescriber both (i) on the extent of Gram-positive coverage needed and will aim to reduce unnecessary use of broad Gram-positive agents such as vancomycin^20^, and (ii) on the breadth of Gram-negative coverage needed^5,6^.

The predictive models that inform the Gram-negative algorithm are derived for each institution prior to the study as previously described^5–7,9^, and use multivariable logistic regression modelling incorporating the following common epidemiologic predictors (age, sex, community vs. ward vs. ICU infection onset, prior microbiologic results and antimicrobial exposures, and admitting service)^5,6^. The model generates a predicted probability that a Gram-negative organism would be susceptible to a given antibiotic, and if this probability exceeds 80% for lower risk patients (qSOFA = 0-2) or 80-90% for higher risk patients (qSOFA = 3) ^21^, then that antibiotic can be recommended. In general, the algorithm is designed to suggest the lowest ‘spectrum’ antibiotic on a designated antibiotic cascade, unless there are specific requirements for additional coverage (e.g., the requirement for Gram-positive coverage for a given syndrome, allergy or drug interaction/contraindication/adverse reaction, or for pseudomonal coverage [e.g., for febrile neutropenia, necrotizing fasciitis, otitis externa, bronchiectasis etc.]) or allergy or drug interaction/contraindication/adverse reaction (see Supplemental Figures 1-3). A supplemental narrow spectrum Gram-positive agent (e.g. cefazolin) could also be combined with the Gram-negative focused agents if needed (e.g. as carbapenem sparing), though general preference will be for maintaining monotherapy. If a particular recommendation could possibly, but not definitively, conflict with a reported allergy (e.g., the allergy label is vague) then the recommendation may still be made with the proviso that the allergy characteristics are clarified (i.e., there is no concern for true allergy or drug reaction by the prescribing team) prior to prescribing. It will be communicated that this can be done in conjunction with the ward/unit pharmacist, as would be done typically in the standard process of care when prescribing antibiotics. qSOFA values will be determined from the most recent available vital signs. Use of vasopressors at the time of assessment will be considered equivalent to qSOFA = 3. For each institution, these models are derived from thousands of patients over multiple years and then validated in the time periods pre-dating the intervention to confirm appropriate performance and calibration. The algorithms have been piloted, tested, and validated among consecutive patients with suspected or confirmed infection ^5–7,9^. Predictions and recommendation thresholds may be modified/customized for specific institutions in the presence of institution-specific underreporting of antibiotic susceptibility (e.g., ceftriaxone) for certain bacterial genus/species. Moreover, a tolerance of 1-2% around thresholds will be accepted (e.g., susceptibility of 79% can be considered approximately 80%). The final algorithm recommendations will be communicated to the most responsible physician and/or prescribing team via a chart note. If there is a recommended change to the patient’s antibiotic therapy, it will be communicated directly to the team (e.g., page, electronic message).

The control period will involve no intervention (i.e., no recommendation made and no communication to the care team) and will thus remain as usual care, i.e., the typical care that is provided to patients with suspected infection at the respective institution and by their usual care providers.

We do not expect there to be significant carry-over effects of the intervention to control periods nor do we expect there to be contamination across clusters. This is because patients will not be included in both periods, and this intervention is unlikely to durably change provider’s behaviours. There are no planned wash out periods. There will be a single lead pharmacist at each site who will be communicating the recommendations (see Supplemental Materials for communication template) and they will not be involved in the routine care of patients at this empiric window, other than those in the intervention group.

### Recruitment

The study intervention is directed towards medical, surgical, and critical care services, with their associated acute care providers who are managing inpatient (admitted) adult patients with suspected sepsis. We will recruit hospital services and associated providers at the start of the trial, through permission from the medical directors (gatekeepers) responsible for these services. We will ask the medical directors to disseminate an information sheet to their physicians in advance of the trial. This information sheet indicates that providers within these services may receive prescribing advice which they are free to implement or ignore. Moreover, on the communication of prescribing advice, we will provide a description of the study and clearly indicate that the decision to consider the advice is at the discretion of the prescriber, and that they may choose not to use this information/participate. Further details on provider and patient consent regarding the intervention and data collection are discussed in the ETHICS section. There will be minimal recruitment time lag, and we will attempt to initiate sites and clusters simultaneously, where possible, and ideally within 3-months of each other. We have estimated that within each cluster, there will be an average of 40 eligible patients per 3-month (13-week) unit, based on prior data ^5,20^. We will be collecting data on the outcomes of the patients that are cared for by the providers working within the intervention/control periods who were deemed eligible.

### Primary Outcome

1. Antibiotic de-escalation from initial empiric index regimen at 48 hours from receipt of antibiotics (index). We define de-escalation as the cessation of a broad-spectrum Gram-positive active agent (i.e. vancomycin, daptomycin, clindamycin, or linezolid), and/or the change of a broad-spectrum Gram-negative active agent to a narrower antibiotic, as specified in an a priori cascade favouring narrow over broad (ciprofloxacin < ceftriaxone < ceftazidime < piperacillin-tazobactam < meropenem). See Supplemental Materials for complete de-escalation cascade which considers all possible antibiotics (excluding atypical agents including macrolides, doxycycline/tetracycline, and metronidazole). Complete cessation of Gram-negative therapy will also be considered de-escalation.

### Secondary Outcomes – Clinical

1. Time to adequate therapy for patients with positive blood cultures (hours from time of collection of first index blood culture collection (regardless of positivity) to first dose of agent(s) active against all pathogen(s) in the peri-index positive blood cultures). Antibiotics given within 24-hours prior to the first index blood collection will be considered and assigned a time of 0 hours if they were adequate.
2. Receipt of adequate antibiotic therapy within 48 hours (or discharge if earlier) from first index blood culture collection for patients with positive blood cultures (active against all pathogens in positive peri-index blood cultures).
3. Receipt of adequate antibiotic therapy within 48 hours (or discharge if earlier) from first index blood culture collection for patients with positive non-screening index cultures including blood (active against all pathogens in peri-index positive cultures).
4. In-hospital mortality, during index admission, and within 90 days of index.
5. Hospital length of stay on index admission (days) up to 90 days.
6. Extent of antibiotic de-escalation at 48 hours from index or discharge if earlier (ordinal value up or down the de-escalation cascade, which is additive in the event of both Gram-positive and Gram-negative de-escalation, see Supplemental Materials for the full cascade).
7. Antibiotic spectrum rank at 7 days from index antibiotic treatment (or at discharge if earlier).
8. Positive stool testing for *Clostridioides difficile* during index admission, and within 90 days of index.
9. New requirement for dialysis during index admission, and within 90 days of index.
10. Total antibiotic days of therapy (DOT) during the first 7 days from index antibiotics (or prior to discharge if earlier), including by spectrum-level.
11. Newly started Gram-positive or Gram-negative antibiotic coverage, or increases in spectrum of antibiotic therapy, as part of the intervention recommendation.
12. Time to de-escalation (hours from receipt of index antibiotics).

### Secondary Outcomes – Process

1. Recommended change in antibiotic therapy by Gram-negative model.
2. Recommended change in antibiotic therapy by Gram-negative model was accepted (acceptance defined as change to some or all of antibiotic therapy as recommended within 24 hours of index antibiotics).
3. Recommended change in antibiotic therapy by Gram-positive algorithm.
4. Recommended change in antibiotic therapy by Gram-positive algorithm was accepted (acceptance defined as change to some or all of antibiotic therapy as recommended within 24 hours if index antibiotics).

### Secondary Outcomes – Safety

1. Escalation of antibiotic therapy (apart from recommended escalation) within 7 days (or prior to discharge if earlier) of index antibiotics (defined as the addition of a concurrent Gram-positive antibiotic to the existing therapy or the increase of spectrum of a Gram-negative active antibiotic). Peri-operative antibiotics will not be considered escalation.
2. Admitted to ICU at day 7 or not alive at day 7 from time of index antibiotics.
3. Time to adequate therapy for patients with positive non-screening index cultures including blood cultures (hours from time of collection of first index blood culture collection (regardless of positivity) to first dose of agent(s) active against all pathogen(s) in the positive peri-index non-screening cultures). Antibiotics given within 24-hours prior to the first index blood culture collection will be considered and assigned a time of 0 hours if they were adequate.

When considering days from index, the days from index is equivalent to index + 7 (e.g., if index day is January 1st, then 7 days from index is January 8^th^). Discharge is defined as: Discharge from hospital, transfer out of acute care, or death. Peri-index cultures are defined as those collected within -72 hours to +12 hours from first dose of index antibiotics. Common blood culture contaminants (including but not limited to *Corynebacterium sp*., coagulase negative staphylococci other than *S*. *lugdenensis*, *Bacillus sp*, and diphtheroids) will not be considered in the determination of adequate therapy. Adequacy outcomes will also be stratified by AmpC and non-AmpC producing organism containing positive cultures.

### Randomization

All 18 participating clusters (six at each of three institutions) will be randomized once-off at study initiation to one of the two sequences. Institutions will be started either simultaneously or in short sequence. An independent (i.e., not familiar with the sites) statistician will generate the allocation sequence using a computer-generated allocation process (concealed from site investigators and analysts), stratified by institution and service to facilitate balance of intervention and control clusters within hospitals and support study feasibility (i.e., not having all clusters receiving the intervention simultaneously at a given hospital) and using a permuted block design with length 2.

### Blinding and Bias Mitigation

Our intervention is fundamentally provider decision support, and we cannot achieve blinding for the physician or the study team (e.g., the pharmacist who provides the prescribing advice). We will mitigate selection bias through identifying eligible patients during intervention and control periods in a pre-specified and protocolized fashion using objective criteria.

### Intervention Administration

A stewardship pharmacist at each participating institution will run the day-to-day trial operations. They will collect data on eligible patients within each cluster, and they will provide the intervention to the prescribers working within the intervention clusters. It is expected that each pharmacist will identify around 3-4 eligible patients and their providers to manage per day, and the remaining time will be used to prospectively collect data on existing patients in intervention and control clusters. A lead pharmacist will help coordinate activities of all three pharmacists, maintain weekly virtual meetings across sites, and ensure the execution of our data management plan. The principal investigator at each site will help with any issues that arise with the research pharmacist at their given site and will adjudicate outcomes.

### Intervention Workflow

Each day the study pharmacist uses their local electronic medical records to identify patients (in both intervention and control clusters) newly started on eligible antibiotics within the last 24 hours. They will then identify patients who have had blood cultures ordered within 12 hours before or after antibiotic initiation. For those patients in intervention clusters meeting the full eligibility criteria, the pharmacist will then review their prior microbiology and epidemiologic risk factors for antimicrobial resistance, input into resistance probability calculators, and will make a recommendation to the MRP regarding either changing antimicrobial therapy (to a narrower, or sometimes broader, antibiotic regimen), or no recommended change based on the predefined algorithms (Figure 1). Other relevant involved teams, including infectious diseases, may be included on these communications. Some recommendation thresholds/predictions may be modified based on local microbiology practices (e.g,. historical censoring of beta-lactam susceptibilities in AmpC species). The recommendation will be summarized in an antimicrobial stewardship study note within the patient’s chart (see sample note in Supplemental Materials). The study pharmacist will record whether the advice was accepted and whether orders were changed (if a change is recommended). Acceptance of recommendation will be defined as a change to some or all recommendation regimens within 24 hours from the index time. For those patients meeting full eligibility criteria in the control periods, the study pharmacist will include them for data collection but will not provide advice or contact the provider. Individuals screened but not eligible for intervention will have their reason for exclusion recorded for the purpose of generating a CONSORT flow diagram.

**Figure 1:**
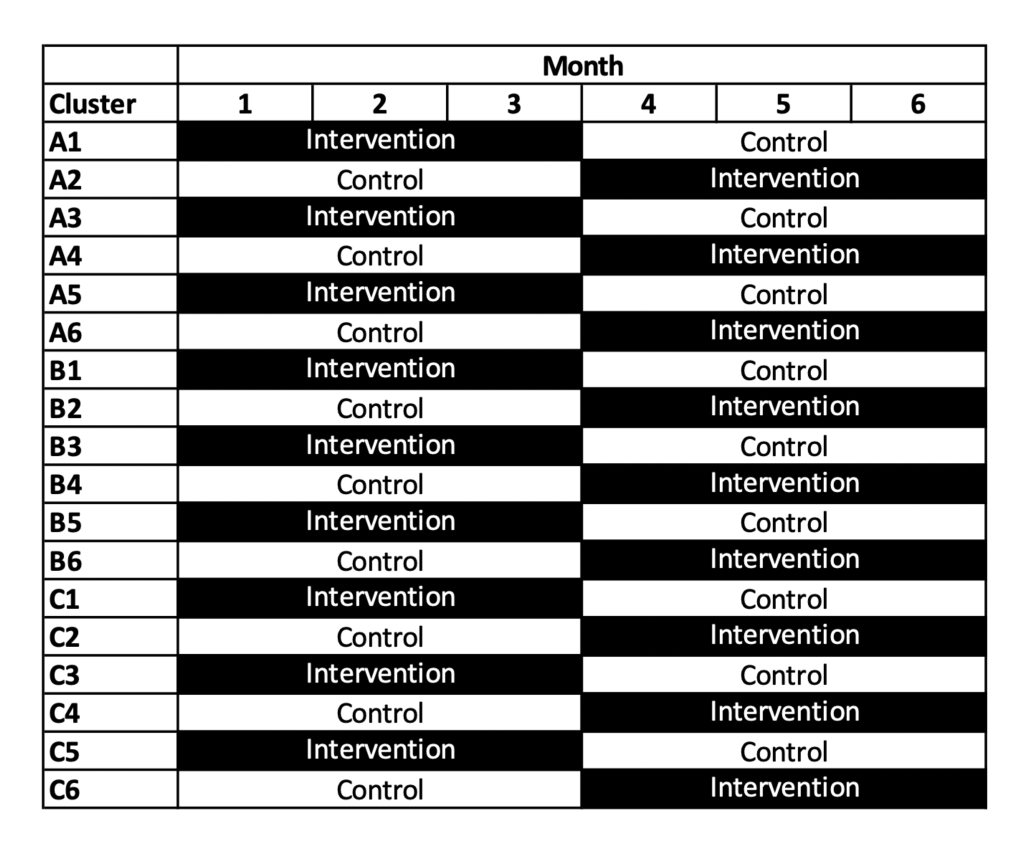
18 trial clusters (across three institutions - A, B, and C) with two cross-over periods.

### Potential Risks and Benefits

The cluster-level intervention will be occurring at the hospital service/unit level, giving providers support for their empiric antibiotic prescribing in sepsis. There is no risk for the safety of providers. For patients being cared for by the providers, there is minimal risk of the intervention as we will be providing only treatment options to the provider, who will retain the autonomy and authority to select a treatment regimen that they believe to be safe and effective for the patient. Patient outcomes from our prior quasi-experimental studies have supported the safety of these approaches^5,20^. Moreover, traditional antibiotic stewardship audit and feedback interventions administered to prescribers have well demonstrated safety in critically ill patients^22^. Patients of providers who receive the prescribing guidance stand to benefit from reduced use of broad-spectrum antimicrobial therapy during their hospital admission, which may result in reduced drug complications (e.g. acute kidney injury), reduced gut flora disruption (e.g., CDI), and reduced risk of antimicrobial resistant organism (ARO) colonization.

### Follow-Up Data Collection

For eligible patients in the intervention and control periods, study pharmacists will perform ongoing data collection for the same admission, up to 90 days from index event. While expected to be rare, should the pharmacist notice a clear discrepancy in treatment and microbiology when reviewing outcomes that could impact clinical care, the care team will be notified. Study data are collected via routinely collected sources (EMR) and will be stored in a de-identified fashion. Data will be collected and stored in a remotely accessible, secure, data entry platform. This data platform will contain relevant covariates and outcome fields, as outlined in our case report form. Study data are de-identified routinely. At the time of entry into the data platform, patients will be assigned a unique study ID and this will be kept separate in a master linking file.

### Sample Size Calculation

We calculated the target sample size for our trial using an open-source application^23^. A total of 18 clusters in a two-period two-sequence cross-sectional cluster cross-over design with an average of 40 patients per cluster-period will achieve 80% power at two-sided *α* =0.05 to detect a clinically important absolute difference of 7.5% (or 37.5% relative improvement) in the primary outcome (de-escalation within 48 hours), assuming a control period proportion of 0.20, a within-period intracluster correlation coefficient (ICC) (calculated from preliminary data) of 0.032, a cluster autocorrelation coefficient (CAC) of 0.86 and a coefficient of cluster size variation of 100% ^5^. We chose a 7.5% absolute improvement as a clinically relevant change and based upon plausible expected impacts of the intervention (including prescribers that don’t follow the recommendations) from prior pilot work which showed effect sizes of similar magnitudes ^5^. Given that this is a cluster randomized trial, with the cluster assignments specified in advance, cluster size heterogeneity will be present, and the final sample size may be over or under the target of 1,440 patients.

### Data Analysis

All analyses will be conducted according to the intention-to-treat principle. The intervention is delivered to providers at the hospital ward/unit level, but the unit of analysis will be the individual patients. The primary outcome will be analyzed using generalized linear mixed effects regression with binomial distribution and log link to produce the intervention effect as a risk ratio. In case of non-convergence, Poisson regression with robust standard errors will be used. The treatment effect will be expressed as risk ratio with 95% confidence interval. The model will include fixed effects for treatment and period to account for the cross-over design, as well as fixed effects for the stratification factors (site and service) and the following a priori patient risk factors to improve power and efficiency: age, sex, comorbidity index, and severity of illness (qSOFA score). We will account for intracluster correlation in our model through inclusion of cluster and cluster-period random effects to allow for a different within and between period intracluster correlation coefficient ^24^. The Kenward-Roger correction will be applied to minimize small sample bias. Secondary outcomes will be analyzed using a similar approach, with binomial distribution for dichotomous variables (e.g. mortality), Poisson or negative binomial distribution for count variables (e.g. antibiotic therapy days), proportional odds model for ordinal outcomes (e.g. extent of antibiotic de-escalation), and normal distribution (with log transformation in the case of skewness) for continuous variables (e.g. hospital length of stay).

If it is necessary to stop the trial early, before all clusters have completed their cross-over, then separate analyses will be performed^25^. In particular, sites that have completed their cross-over (i.e., sites with complete data for both periods) will be analyzed as a two-period cluster cross-over trial as described above, whereas sites that have completed only the first period will be analyzed as a parallel arm cluster randomized trial^25^. Thus, at these sites a similar statistical model will be used but adapted to a standard two-arm parallel design (without a period effect). Treatment effect estimates and standard errors from the two analyses will be pooled to obtain an overall estimate using fixed-effects meta-analysis with inverse variance weights. All primary and secondary outcomes will be analyzed using a similar approach.

This study has been registered at clinicaltrials.gov and will be reported in line with the CONSORT extension statement for cluster randomized cross-over trials ^26^.

### Safety Outcomes

We will be collecting safety indices, including admission to ICU or mortality at day 7 from index antibiotics as well as escalation of antimicrobial therapy (apart from those recommended) as part of our planned trial outcomes. These will be reviewed at the completion of the trial by the data safety monitoring board (DSMB).

### Trial Steering Committee and DSMB

We will operate a trial steering committee composed of the co-investigators listed in this protocol, representing all participating sites, that will oversee study operations. We have convened a formal independent DSMB, that will review the trial for any operational, scientific, or safety outcome concerns at the start and completion of the trial (given it is a short duration pragmatic study). Our intent is to initiate all sites and clusters simultaneously or in close succession, with study intervention completion at up to 12-months if lagged initiation. Thus the DSMB would meet prior to the full study initiation, and then after completion which would be 12-15 months (including 3-month follow up period) depending upon whether there is an initiation lag. The DSMB would be asked to meet prior to completion if any safety or feasibility concerns were to arise. Our DSMB will comprise 3-4 external members (not involved in this trial or related publications) and include at least 2 clinical experts in the field and at least 1 biostatistician with expertise in cluster randomized designs. During the trial, the steering committee will have the option to request an interim analysis of the data available to date including safety outcomes (without formal hypothesis testing/stopping criteria) and potentially primary outcome if requested, with review by the DSMB, and this would include if the trial completion (including follow up) is expected to exceed 12-months from study start. The details of this review will be outlined in the DSMB charter. Any DSMB recommendations would be communicated with the trial steering committee and shared with the REB.

## ETHICS

The Ottawa Statement on the Ethical Design and Conduct of Cluster Randomized Trials provides 15 recommendations across seven domains to address the complex ethical issues raised in these types of trial designs^27^. We adhere to its recommendations as follows.

### Justifying the cluster randomized design

Cluster randomization was chosen over individual randomization: (1) to help prevent contamination within services; (2) to facilitate delivery of the intervention in an efficient and effective manner; and (3) to accommodate the presence of trainees which are directly involved in prescribing and associated (by definition) with staff physicians/services. A cross-over design was chosen over a parallel-arm design to (1) improve power and efficiency given the limited number of sites that could feasibly be enrolled and (2) promote a balance in patient case mix across different treatment conditions.

### Research ethics committee review

We have obtained research ethics board approval from all participating sites.

### Identifying research participants

Providers are the research participants as they are the target of the cluster-level intervention (i.e., providers either receive pharmacists’ advice or not, based upon the service they are working under). Patients, however, are not the research participants for two reasons. First, although providers receive an intervention aimed at improving prescribing practices, the provider is expected to act in the best interests of their patients and in accordance with professional practice standards. Second, there is no interaction with patients for data collection, and data will be stored in a de-identified fashion.

### Obtaining informed consent

Informed consent for the study intervention and data collection are separable in cluster trials and should correspond to the participant’s involvement in the study. For providers (the targets of the cluster-level intervention), a modified consent process was obtained for the study intervention: after gatekeeper permission for enrollment of clusters was obtained, providers were notified of the study through multiple mechanisms (e.g., via information sheets shared in advance by medical directors, reminders at the time of the intervention advice) and providers were informed that they are not required to follow the advice given. The rationale for this modification is as follows: (1) receiving feedback on prescribing practices via pharmacist’s recommendation poses minimal risk and providers retain autonomy to make their prescribing decision, (2) obtaining individual informed consent from all providers is considered infeasible because of the large number of providers, and because of changes in providers associated with a given service and cluster), and (3) providers are made aware of the study through multiple approaches and will be aware that they are free to ignore the advice given. Patient outcomes related to the provider intervention will be collected, and although we don’t consider patients as the trial participants, a waiver of consent for the use of patient-level data was obtained given that (1) the data collection is low risk and unlikely to impact the welfare of participants, and (2) it is impractical to collect individual patient consent for data collection.

### Gatekeepers

We will seek approval from medical directors responsible for the enrollment and randomization of their cluster(s).

### Assessing benefits and harms

Existing evidence demonstrating safety and potential for benefit are explicated in the background section. Patients in the control arm receive usual care and are thus not deprived of effective care. The risks associated with data collection are minimal, and data is stored in a de-identified fashion.

### Protecting vulnerable participants

To protect the confidentiality of individuals, study data will be stored in de-identified fashion as previously described.

## DISSEMINATION

Research findings will be published in international open-access peer-reviewed journal(s) and presented at meetings or conferences in abstracts, poster or oral presentations. The results of the study will be reported as per the SPIRIT guidelines for RCTs^28^. Authorship will be assigned in accordance with the International Committee of Medical Journal Editors recommendations.

## PARTICIPANT ENGAGEMENT

We engaged with provider clinical leads to obtain permission to conduct the study on their units with their prescribers. In addition, providers received communications in advance, and at the time of the intervention. We incorporated feedback from providers in advance of the study during orientation sessions explaining the nature of the study.

## Data Availability

This manuscript is a study protocol - there are no data presented herein.

## AUTHORS CONTRIBUTIONS

DM, ND, ME, CG, MT, CEG, and CN contributed to study conceptualization and design. All authors contributed to the development and critical review of the protocol manuscript.

## FUNDING STATEMENT

This work was supported by the Canadian Institutes of Health Research (Project Grant # 202203ARG-482485-RC1-ADYP-253713).

## COMPETING INTERESTS STATEMENT

All authors have no relevant conflicts of interest to declare.

## SUPPLEMENTAL FIGURES

**Supplemental Figure 1:**
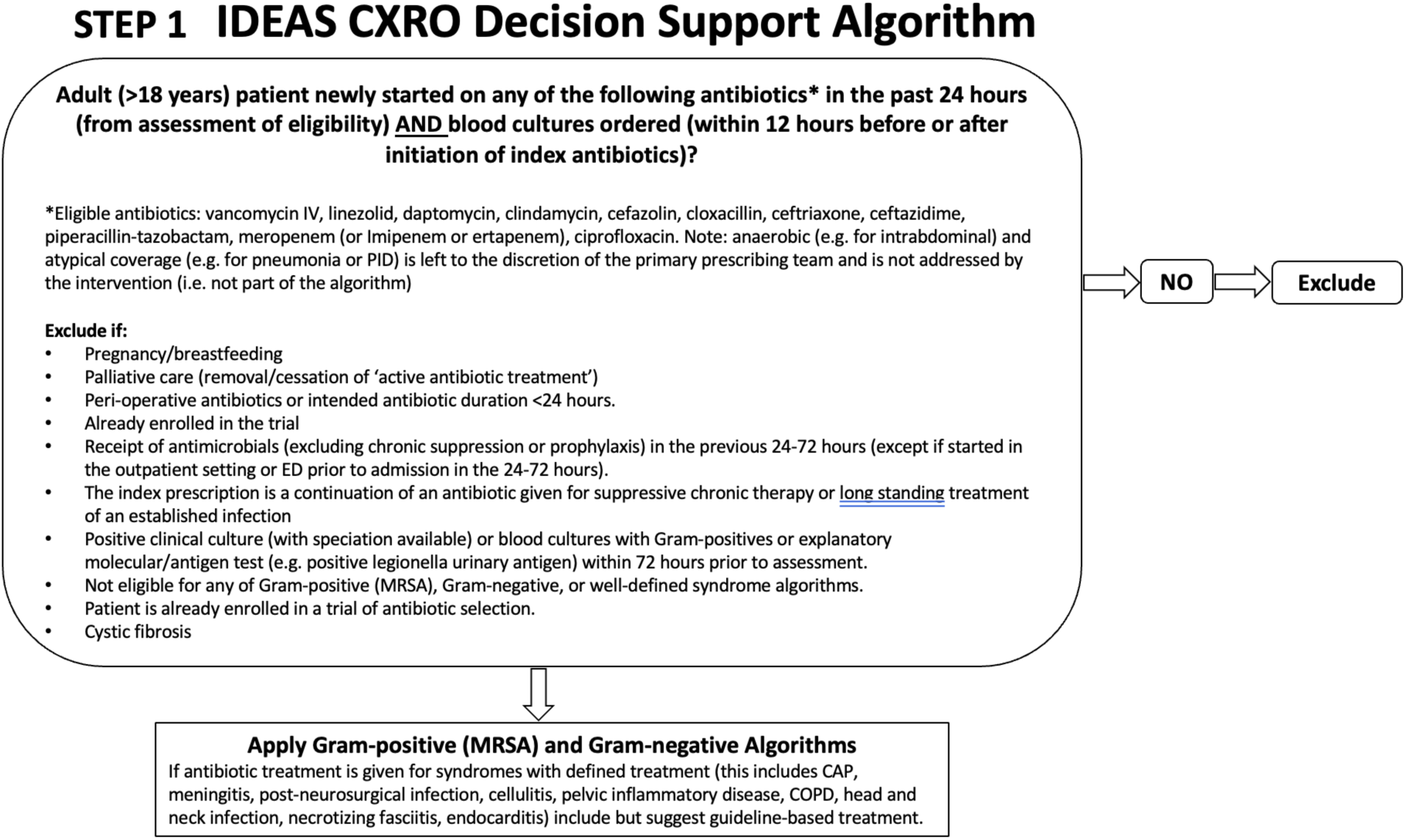
Eligibility.

**Supplemental Figure 2:**
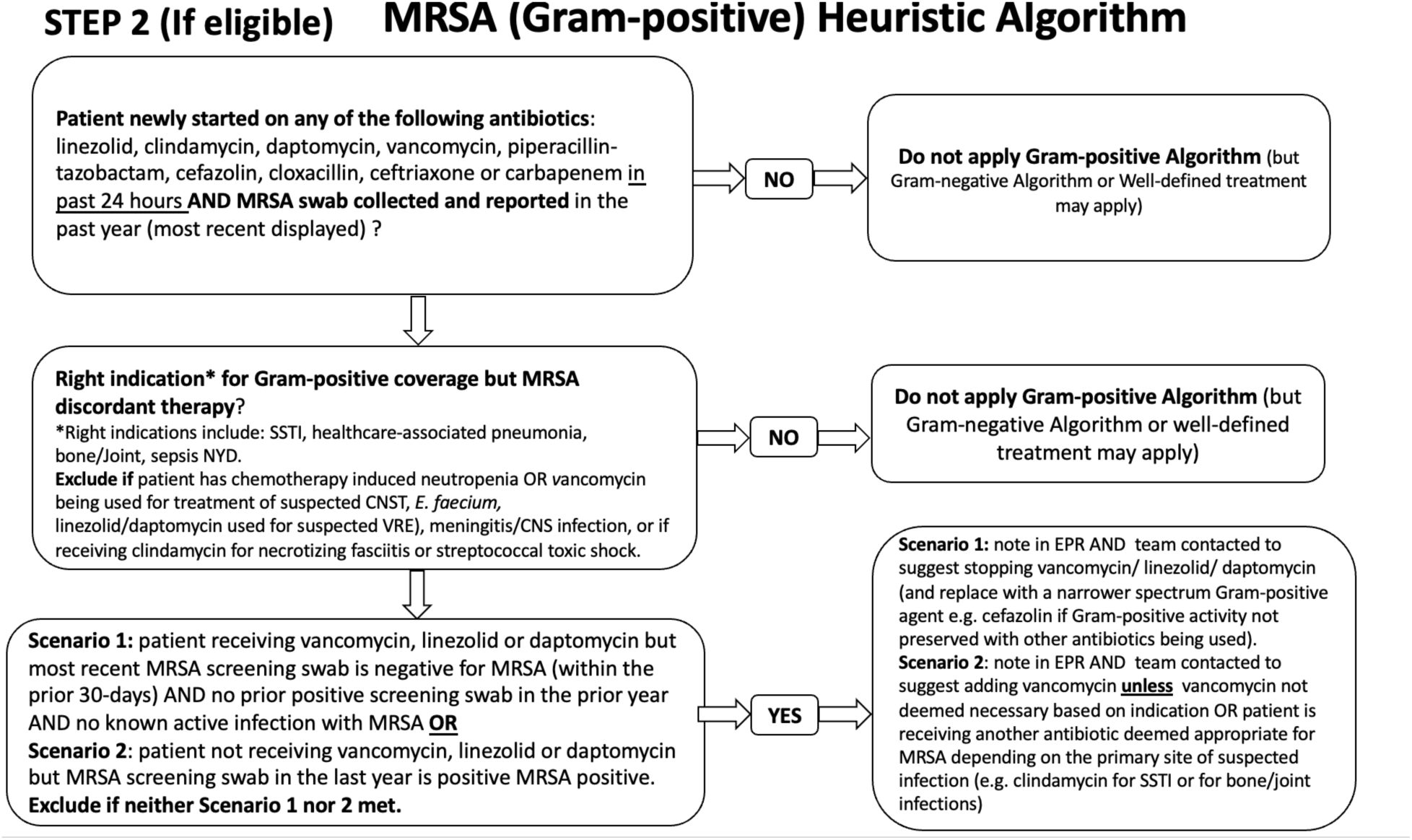
Gram-positive Algorithm.

**Supplemental Figure 3:**
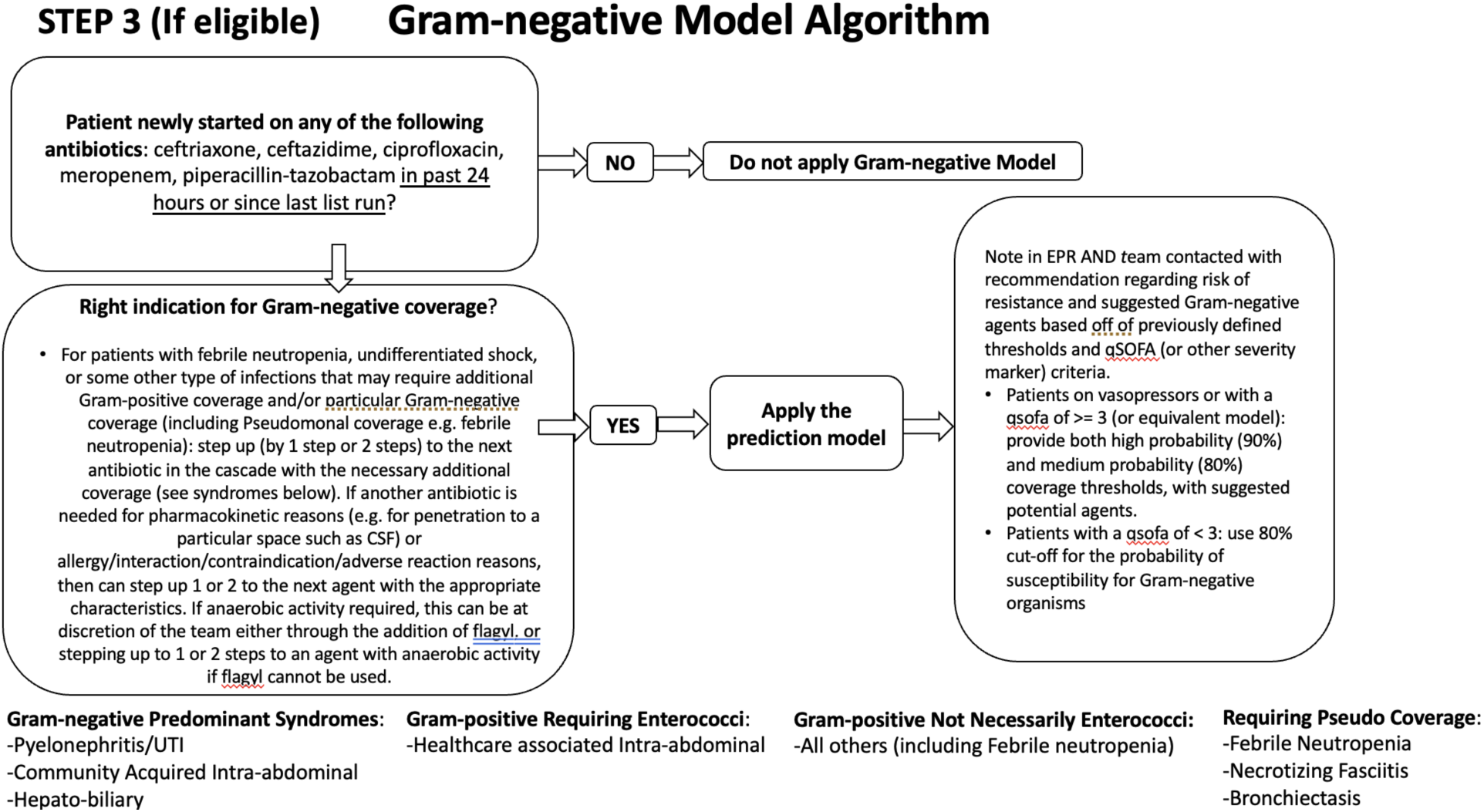
Gram-negative Algorithm.

## SUPPLEMENTAL MATERIALS

### EXAMPLE OF EMR REPORT OF PRESCRIBING ADVICE

#### IDEAS-CRXO Study Note

We are providing antibiotic recommendations as part of the IDEAS-CRXO study. The study provides recommendations to prescribers to support **empiric antibiotic selection**. It is based on a treatment algorithm that takes into consideration patient demographics, prior antibiotic therapy, and prior culture results from the hospital chart.

#### IDEAS-CRXO Study Assessment

**<u>[MRSA Coverage]</u>**

Your patient is receiving [name of antibiotic] and previous [cultures or MRSA screening swabs] have demonstrated [presence/absence of MRSA]. [If present: MRSA colonization is a risk factor for MRSA infections when identified within the last year] OR [If absent: Lack of MRSA colonization within the prior 30 days significantly reduces the risk of MRSA infection] [Butler-Laporte G, et al. Antimicrob Agents Chemother 60:7444–48.].

#### Study Recommendation

As patient had MRSA identified within the last year, [current antimicrobial therapy is appropriate (if on vancomycin) OR suggest changing to/adding vancomycin (or another appropriate antibiotic).]

OR

As patient has had a negative MRSA screening swab within the prior 30 days, please consider [stopping vancomycin OR changing to a beta-lactam (specify)].

<u>[Syndrome-based recommendation]</u>

Your patient is currently receiving [name of antibiotic(s)] for [cellulitis OR community-acquired pneumonia OR COPD exacerbation OR community-acquired meningitis OR post-neurosurgical CNS infection].

#### Study Recommendation

Based on the hospital’s local empiric therapy guidelines, [current antimicrobial therapy is appropriate.] OR [consider changing current antibiotic regimen to [GL concordant therapy].

<u>[Gram-negative Coverage]</u>

**[If qSOFA score ≤ 2]**

You are currently providing your patient with Gram-negative antibiotic coverage for their suspected infection. Based on epidemiologic models*, the following are the <u>predicted</u> probabilities of susceptibility of Gram-negative organisms for various antibiotics: Low Probability of Activity (<80%): Ciprofloxacin

Moderate to High Probability of Activity (≥80%): Ceftriaxone, Ceftazidime, Pip-Tazo, or Meropenem

*As your patient’s infection severity is considered to be low based on a qSOFA score of ≤ 2, a threshold of at least 80% is considered acceptable. Probabilities that meet the threshold are in green and probabilities that are less than the threshold are in red. [Cressman AM et al. Clin Infect Dis 2019;69(6):930–7.]*

[Select one of the following]

[If adequate coverage]

Based on the above probabilities, the antibiotic the patient is currently receiving ([name of antibiotic]) is predicted to provide adequate coverage of Gram-negatives in their suspected infection.

#### Study Recommendation

Current antimicrobial therapy is appropriate.

[If adequate coverage but to narrow spectrum]

Based on the above probabilities, the antibiotic the patient is currently receiving ([name of antibiotic]) is predicted to provide adequate coverage of Gram-negatives in their suspected infection, however, a narrower spectrum agent(s) ([name of antibiotic]) should also provide adequate coverage.

#### Study Recommendation

please consider changing [name of current antibiotic] to [name of antibiotic] [add indication if need to step up for gram positive or anaerobic coverage e.g. pneumonia].

[If inadequate coverage]

Based on the above probabilities, the antibiotic the patient is currently receiving ([name of antibiotic]) is predicted to provide a low probability of coverage of Gram-negatives in their suspected infection.

#### Study Recommendation

please consider changing [name of current antibiotic] to [name of antibiotic]. [add indication if need to step up for gram positive or anaerobic coverage e.g. pneumonia].

[If model does not identify any antibiotic with ≥ 80% threshold—**Note**: do NOT provide antibiotic probabilities]

Based on the predictive model, the patient is at high risk of a multi-drug resistant gram-negative organism.

#### Study Recommendation

Consider use of combination therapy and/or consult the Infectious Diseases Service for advice.

#### [If qSOFA score is 3 or on vasopressor]

You are currently providing your patient with Gram-negative antibiotic coverage for their suspected infection. Based on epidemiologic models*, the following are the <u>predicted</u> *probabilities of susceptibility* of Gram-negative organisms for various antibiotics:

Low Probability of Activity (<80%): Ciprofloxacin

Moderate Probability of Activity (80-89%): Ceftriaxone, Ceftazidime, or Pip-Tazo

High Probability of Activity (≥ 90%): Meropenem

*Generally a threshold of at least 80% is recommended. Your patient’s infection severity is considered to be high based on [a qSOFA score of 3 OR the need for vasopressor support] and a higher threshold may be considered. Probabilities ≥ 90% are in green, between 80-90% are in orange, and below the 80% threshold are in red. [Cressman AM et al. Clin Infect Dis 2019;69(6):930–7.]*

[Select one of the following]

[If current antibiotic meets ≥90% threshold]

Based on the above probabilities the antibiotic the patient is currently receiving ([name of antibiotic]) is predicted to provide a high probability of coverage of Gram-negatives in their suspected infection.

#### Study Recommendation

Current antimicrobial therapy is appropriate.

[If adequate coverage (meets ≥90%) but to narrow spectrum]

Based on the above probabilities, the antibiotic the patient is currently receiving ([name of antibiotic]) is predicted to provide adequate coverage of Gram-negatives in their suspected infection, however, a narrower spectrum agent(s) ([name of antibiotic]) should also provide adequate coverage.

#### Study Recommendation

please consider changing [name of current antibiotic] to [name of antibiotic] [add indication if need to step up for gram positive or anaerobic coverage e.g. pneumonia].

[If current antibiotic does not meet 80% threshold]

Based on the above probabilities the antibiotic the patient is currently receiving ([name of antibiotic]) is predicted to provide a low probability of coverage of Gram-negatives in their suspected infection.

#### Study Recommendation

Please consider selecting an alternate agent with at least moderate (∼80-90%) or high (>90%) predicted probability of Gram-negative activity as provided above. Please consider the need for Gram-positive activity if selecting an alternative agent.

[If current antibiotic is between 80-90% threshold and an antibiotic with >= 90% probability exists]

Based on the above probabilities the antibiotic the patient is currently receiving ([name of antibiotic]) provides a moderate probability of coverage of Gram-negatives in their suspected infection.

#### Study Recommendation

If clinically indicated, consider selecting an antibiotic with a ≥ 90% predicted probability of Gram-negative activity as provided above. Please consider the need for Gram-positive activity if selecting an alternative agent.

[If current antibiotic is between 80-90% threshold with no ≥90% probability options]

Based on the above probabilities the antibiotic the patient is currently receiving ([name of antibiotic]) provides a moderate probability of coverage of Gram-negatives in their suspected infection.

Study Recommendation: Continue [name of antibiotic]. If obtaining ≥90% probability is clinically indicated, consider use of combination therapy and/or consult the Infectious Diseases Service for advice.

[If model does not identify any antibiotic with ≥ 80% threshold—**Note**: do NOT provide antibiotic probabilities]

Based on the predictive model, the patient is at high risk of a multi-drug resistant gram-negative organism.

#### Study Recommendation

Consider use of combination therapy and/or consult the Infectious Diseases Service for advice.

*NOTE: The advice is not a replacement for clinician judgement and is only provided at this single point in time (no ongoing management advice). Models are based on data from Microbiology/Pharmacy systems and not from patient history. Providers **are not obligated** to follow this advice and should regularly reassess antibiotic treatment based on ongoing clinical status and/or microbiologic/diagnostic results as per usual patient care. Antibiotics within 1-2% of a threshold were assumed equivalent to that threshold.*

*[AS APPLICABLE] The predicted probabilities of have been adjusted (+ %) due to historic under-reporting of beta-lactam susceptibilities.*

*The antimicrobial stewardship pharmacist is available at [pager] for dosing/prescribing support. For further information on the study, please visit the intranet site (INSERT LINK HERE)*

*\*See links for supporting literature for the IDEAS algorithms/models.(<u>MacFadden et al,</u> <u>Bucheeri et al., Elligsen et al.)</u>*

## CASCADE FOR INTERPRETING GRAM-NEGATIVE STEP CHANGES

### HIGHEST

1. Meropenem/imipenem/ertapenem/colisitin/cefiderocol/ceftazidime+avibactam/ tigecycline/ceftolozane+tazobactam/amikacin
2. Piperacillin+tazobactam/IV fosfomycin
3. Ceftazidime/tobramycin/gentamicin/cefepime
4. Ceftriaxone/ceftaroline/cefixime/moxifloxacin/levofloxacin/ amoxicillin+clavulanate
5. Ciprofloxacin
6. Trimethoprim/Sulfamethoxazole, first AND second generation cephalosporins, penicillins, oral fosfomycin, nitrofurantoin
7. Discontinued

### LOWEST

## ELIGIBILITY TIMING AND INTERVENTION STEPS

1. Initial screening (list generated from EMR)
2. Confirmation of eligibility - EMR-based chart review. After eligibility is confirmed from EMR, patient is entered into the database (study ID assigned). Applied to both control and intervention groups. -------- Any patients included after Step 2 would be included in the data collection (and ultimately analyses). However, if found to be ineligible after Step 2, no recommendation is made.
3. Communication of recommendation. If during the process of generating the recommendation (for the intervention patients) and communicating it, the patient is no longer eligible, they will be included for data collection/outcomes but no recommendation will be made. This includes patients who might have antibiotics stopped, for example.

## Notes

### Competing Interest Statement

The authors have declared no competing interest.

### Clinical Trial

NCT06103500

### Funding Statement

This work was funded by the Canadian Institutes of Health Research (Project Grant # 202203ARG-482485-RC1-ADYP-253713). 

### Author Declarations

Clinical trails Ontario (CTO) gave province-wide ethics approval for this study to be conducted. Clinical trials Ontario (CTO) gave ethical approval for all individual study sites (i.e. Ottawa, Toronto, Mississauga) for this study to be conducted.

